# Covid-19 by Race and Ethnicity: A National Cohort Study of 6 Million United States Veterans

**DOI:** 10.1101/2020.05.12.20099135

**Authors:** Christopher T. Rentsch, Farah Kidwai-Khan, Janet P. Tate, Lesley S. Park, Joseph T. King, Melissa Skanderson, Ronald G. Hauser, Anna Schultze, Christopher I. Jarvis, Mark Holodniy, Vincent Lo Re, Kathleen M. Akgün, Kristina Crothers, Tamar H. Taddei, Matthew S. Freiberg, Amy C. Justice

## Abstract

**Background:** There is growing concern that racial and ethnic minority communities around the world are experiencing a disproportionate burden of morbidity and mortality from symptomatic SARS-Cov-2 infection or coronavirus disease 2019 (Covid-19). Most studies investigating racial and ethnic disparities to date have focused on hospitalized patients or have not characterized who received testing or those who tested positive for Covid-19.

**Objective:** To compare patterns of testing and test results for coronavirus 2019 (Covid-19) and subsequent mortality by race and ethnicity in the largest integrated healthcare system in the United States.

**Design:** Retrospective cohort study.

**Setting:** United States Department of Veterans Affairs (VA).

**Participants:** 5,834,543 individuals in care, among whom 62,098 were tested and 5,630 tested positive for Covid-19 between February 8 and May 4, 2020.

**Exposures:** Self-reported race/ethnicity.

**Main outcome measures:** We evaluated associations between race/ethnicity and receipt of Covid-19 testing, a positive test result, and 30-day mortality, accounting for a wide range of demographic and clinical risk factors including comorbid conditions, site of care, and urban versus rural residence.

**Results:** Among all individuals in care, 74% were non-Hispanic white (white), 19% non-Hispanic black (black), and 7% Hispanic. Compared with white individuals, black and Hispanic individuals were more likely to be tested for Covid-19 (tests per 1000: white=9.0, [95% CI 8.9 to 9.1]; black=16.4, [16.2 to 16.7]; and Hispanic=12.2, [11.9 to 12.5]). While individuals from minority backgrounds were more likely to test positive (black vs white: OR 1.96, 95% CI 1.81 to 2.12; Hispanic vs white: OR 1.73, 95% CI 1.53 to 1.96), 30-day mortality did not differ by race/ethnicity (black vs white: OR 0.93, 95% CI 0.64 to 1.33; Hispanic vs white: OR 1.07, 95% CI 0.61 to 1.87).

**Conclusions:** Black and Hispanic individuals are experiencing an excess burden of Covid-19 not entirely explained by underlying medical conditions or where they live or receive care. While there was no observed difference in mortality by race or ethnicity, our findings may underestimate risk in the broader US population as health disparities tend to be reduced in VA.

## Introduction

The United States has the highest number of reported symptomatic severe acute respiratory syndrome coronavirus 2 (SARS-CoV-2) infections and related deaths in the world, accounting for one-third of global totals.^1^ There is growing concern that racial and ethnic minority communities are experiencing a disproportionate burden of morbidity and mortality from symptomatic SARS-Cov-2 infection or coronavirus disease 2019 (Covid-19).^2-5^ One report from Milwaukee County, Wisconsin found that Covid-19 cases were largely clustered in predominately African American areas and that this clustering was independent of income.^6^ The largest study to date on racial and ethnic disparities in the US conducted by the Centers for Disease Control and Prevention (CDC) found that black individuals were overrepresented in a national sample of 580 patients hospitalized with Covid-19.^7^ Thus, racial and ethnic disparities in the incidence of and outcomes from Covid-19 have been deemed an urgent public health research priority.^8^ However, most studies investigating racial and ethnic disparities have focused on hospitalized patients or have not characterized which individuals were tested or those who tested positive for Covid-19.^9-13^ In addition, it is not yet known whether such disparities are driven, at least in part, by differences in underlying health conditions, smoking and alcohol use, geographic location, or urban versus rural residence – essential information if we are to design effective interventions.

The electronic health record database of the Department of Veterans Affairs (VA) offers the single largest national data resource available with the necessary information on system-wide testing and detailed medical histories to examine racial and ethnic disparities in the US. We evaluated associations between race/ethnicity and receipt of Covid-19 testing, a positive test result, and 30-day mortality, each outcome conditional on the previous and accounting for a wide range of demographic and clinical risk factors and geographic site of care through May 4, 2020.

## Methods

### Data Source

The VA is the largest integrated healthcare system in the United States and comprises over 1200 points-of-care nationally including hospitals, medical centers, and community outpatient clinics. All care is recorded in a national electronic health record with daily uploads into the VA Corporate Data Warehouse. Available data include demographics, outpatient and inpatient encounters, diagnoses, smoking and alcohol health behaviors, and pharmacy dispensing records.

This study was approved by the Institutional Review Boards of VA Connecticut Healthcare System and Yale University. It has been granted a waiver of informed consent and is Health Insurance Portability and Accountability Act compliant.

### Sample, Follow-up, and Outcomes

All individuals active in clinical care (defined as having at least one clinical encounter between January 1, 2018 and December 31, 2019 and alive as of January 1, 2020) were included in this analysis. We identified individuals tested for Covid-19 from date of first recorded test on February 8, 2020 through May 4, 2020 by using text searching of laboratory results containing terms consistent with SARS-CoV-2 or Covid-19. Nearly all tests utilized nasopharyngeal swabs, 1% were from other sources. Testing was performed in VA, state public health, and commercial reference laboratories using emergency use authorization approved SARS-CoV-2 assays. We did not include antibody tests in this analysis.

If an individual had more than one test and all were negative we selected the date of the first negative test, otherwise we used the date of the first positive test. Baseline for individuals tested for Covid-19 was defined as the date of specimen collection unless testing occurred during hospitalization, in which case it was defined as date of admission. If the admission began more than 14 days prior to testing, which may indicate hospital-acquired infection, we set baseline to 14 days prior to testing to better capture health status prior to SARS-CoV-2 infection. We examined three outcomes: 1) receipt of Covid-19 testing among all in care, 2) a positive test result among individuals tested for Covid-19, and 3) 30-day mortality among Covid-19 cases. Deaths were captured using inpatient records and VA death registry to capture deaths outside of hospitalization.

### Covariates

We selected previously validated cohort characteristics and those that have been evaluated in prior Covid-19 reports. Demographics included age at baseline, sex, race/ethnicity (non-Hispanic white [white], non-Hispanic black [black], and Hispanic), and rural/urban residence. While the VA cares for individuals of other racial and ethnic backgrounds, there were too few tested for Covid-19 to obtain estimates in adjusted analyses at this time. Residence was defined using geographic information system coding based upon established criteria.^14^

We extracted diagnostic codes for asthma, any cancer, chronic obstructive pulmonary disease (COPD), chronic kidney disease, diabetes mellitus, hypertension, liver disease, vascular disease, and alcohol use disorder (definitions provided in **Supplementary Table S1**). Presence of conditions was determined by one inpatient or two outpatient diagnoses in the two years prior to baseline, except for cancer which was ever prior to baseline. Diagnoses made in the 7 days prior to baseline were not included. We used a validated algorithm to capture smoking status^15^ and alcohol consumption.^16^ We collected pharmacy fills for angiotensin converting enzyme (ACE) inhibitors and angiotensin II receptor blockers (ARBs) and identified individuals with active prescriptions in the 30 days prior to baseline. Missing data for smoking and alcohol consumption only affected 8% of individuals included in multivariable models, thus complete case analysis was performed.

### Statistical Analysis

We calculated number of Covid-19 tests per 1,000 individuals in care and Clopper-Pearson 95% confidence intervals (CIs) by race/ethnicity category. Among those tested for Covid-19, we calculated percent testing positive for Covid-19 and 95% CIs by race/ethnicity category. Logistic regression models were used to estimate age-adjusted and fully adjusted associations between race/ethnicity and testing positive for Covid-19. Fully adjusted models included all pre-specified covariates listed above. We further fitted a model conditioned on site of care among sites with at least five positive tests (which captured 99% of Covid-19 cases) to account for site-specific differences in Covid-19 testing. We repeated this modelling procedure to estimate odds ratios (ORs) and 95% CIs between race/ethnicity and 30-day mortality among those testing positive for Covid-19 on or prior to April 3, 2020 to allow all individuals 30 days of follow-up. In sensitivity analyses, we compared associations with testing positive for Covid-19 over two time periods (February 8 to April 3, 2020 and April 4 to May 4, 2020). We also restricted the outcome for 30-day mortality to inpatient deaths to test robustness of associations found in a model including all deaths. Analyses were performed using SAS version 9.4 (SAS Institute Inc., Cary, NC, USA). R version 3.6.3 was used to map Covid-19 cases nationally.

## Results

There were 5,834,543 individuals active in care prior to the Covid-19 pandemic. Most (91%) were men, 74% were white, 19% were black, and 7% were Hispanic (**Table 1**). Ages ranged from 20-105 years, with 24% less than 50 years, 35% 50-69 years, 28% 70-79, and 13% 70 or more years of age, which was similar by race/ethnicity. Of these, 62,098 (10.6 per 1000 in care) were tested for Covid-19, of whom 62% were white, 29% were black, and 9% were Hispanic. There were 5,630 (9.0%) individuals who tested positive for Covid-19 between February 8 and May 4, 2020, of whom 40% were white, 49% were black, and 11% were Hispanic. While 66% of all individuals in care resided in urban areas, 79% of those tested and 92% of those testing positive for Covid-19 resided in urban areas.

**Table 1.** Characteristics of all individuals in care and those tested for Covid-19 as of May 4, 2020

|  | No. in care | (col %) | No. tested | (col %) | No. tested positive | (col %) |
| --- | --- | --- | --- | --- | --- | --- |
| Sample size, n (%) | 5,834,543 | (100.0) | 62,098 | (100.0) | 5,630 | (100.0) |
| Race/ethnicity |  |  |  |  |  |  |
| White | 4,309,613 | (73.9) | 38,774 | (62.4) | 2,261 | (40.2) |
| Black | 1,089,883 | (18.7) | 17,992 | (29.0) | 2,743 | (48.7) |
| Hispanic | 435,047 | (7.5) | 5,332 | (8.6) | 626 | (11.1) |
| Age, years |  |  |  |  |  |  |
| 20-39 | 842,948 | (14.4) | 7,401 | (11.9) | 590 | (10.5) |
| 40-49 | 577,135 | (9.9) | 6,093 | (9.8) | 538 | (9.6) |
| 50-59 | 840,779 | (14.4) | 10,844 | (17.5) | 1,006 | (17.9) |
| 60-69 | 1,210,960 | (20.8) | 15,579 | (25.1) | 1,365 | (24.2) |
| 70-79 | 1,620,765 | (27.8) | 15,358 | (24.7) | 1,420 | (25.2) |
| ≥80 | 741,956 | (12.7) | 6,823 | (11.0) | 711 | (12.6) |
| Sex |  |  |  |  |  |  |
| Female | 522,738 | (9.0) | 7,192 | (11.6) | 467 | (8.3) |
| Male | 5,311,805 | (91.0) | 54,906 | (88.4) | 5,163 | (91.7) |
| Residence type |  |  |  |  |  |  |
| Rural | 2,002,299 | (34.3) | 13,028 | (21.0) | 476 | (8.5) |
| Urban | 3,832,244 | (65.7) | 49,070 | (79.0) | 5,154 | (91.5) |

Abbreviations: Covid-19, coronavirus disease 2019; col %, column percent

The geographic distribution of all 5,630 Covid-19 cases in the VA were similar to known hotspots in the general population, including Northeastern and Midwestern states, Louisiana, and Colorado (**Figure 1a**). Several facilities performing the highest number of tests also had the highest proportion of black individuals in care, including New York City, Chicago, New Orleans, and Detroit **(Figure 1b)**.

**Figure 1.**
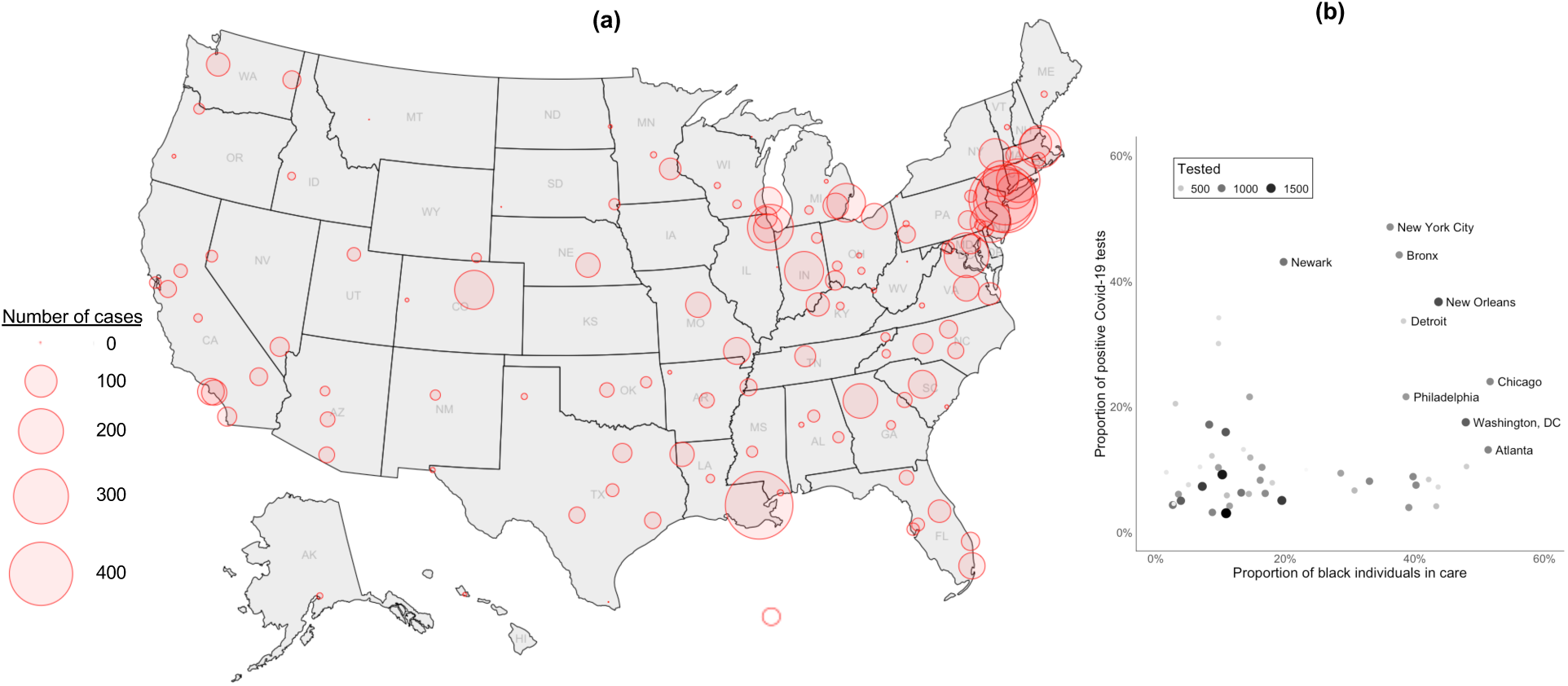
Distribution of 5,630 Covid-19 cases in the US Department of Veterans Affairs as of May 4, 2020 (a) Shown is the distribution of all Covid-19 Veteran cases in the US Department of Veterans Affairs between February 8 and May 4, 2020 and included in the current study. (b) Shown is the proportion of positive Covid-19 test results by the proportion of black individuals care by site among sites with at least 5% positivity, which captures 86% of all Covid-19 cases.

### Rate of Testing and Testing Positive

Testing rates for Covid-19 was higher among black (tests per 1000 in care: 16.4, 95% CI 16.2-16.7) and Hispanic individuals (12.2, 95% CI 11.9-12.5) than white individuals (9.0, 95% CI 8.9-9.1). Testing rates also varied by age, sex, residence type **(Table 2)**. Among white individuals in care, those aged 60-69 years were most likely to be tested (tests per 1000: 13.9, 95% CI 13.6-14.1). However, among black and Hispanic individuals in care, testing rates steadily increased with age with those aged ≥80 years being most likely to be tested (tests per 1000: 52.2, 95% CI 50.4-54.0 for black individuals; 24.3, 95% CI 22.7-26.0 for Hispanic individuals). Among white individuals, women were more likely than men and those residing in urban areas versus rural areas were more likely to be tested for Covid-19. Conversely, among black and Hispanic individuals, men were more likely than women and those residing in rural areas versus urban areas were more likely to be tested for Covid-19.

**Table 2.**
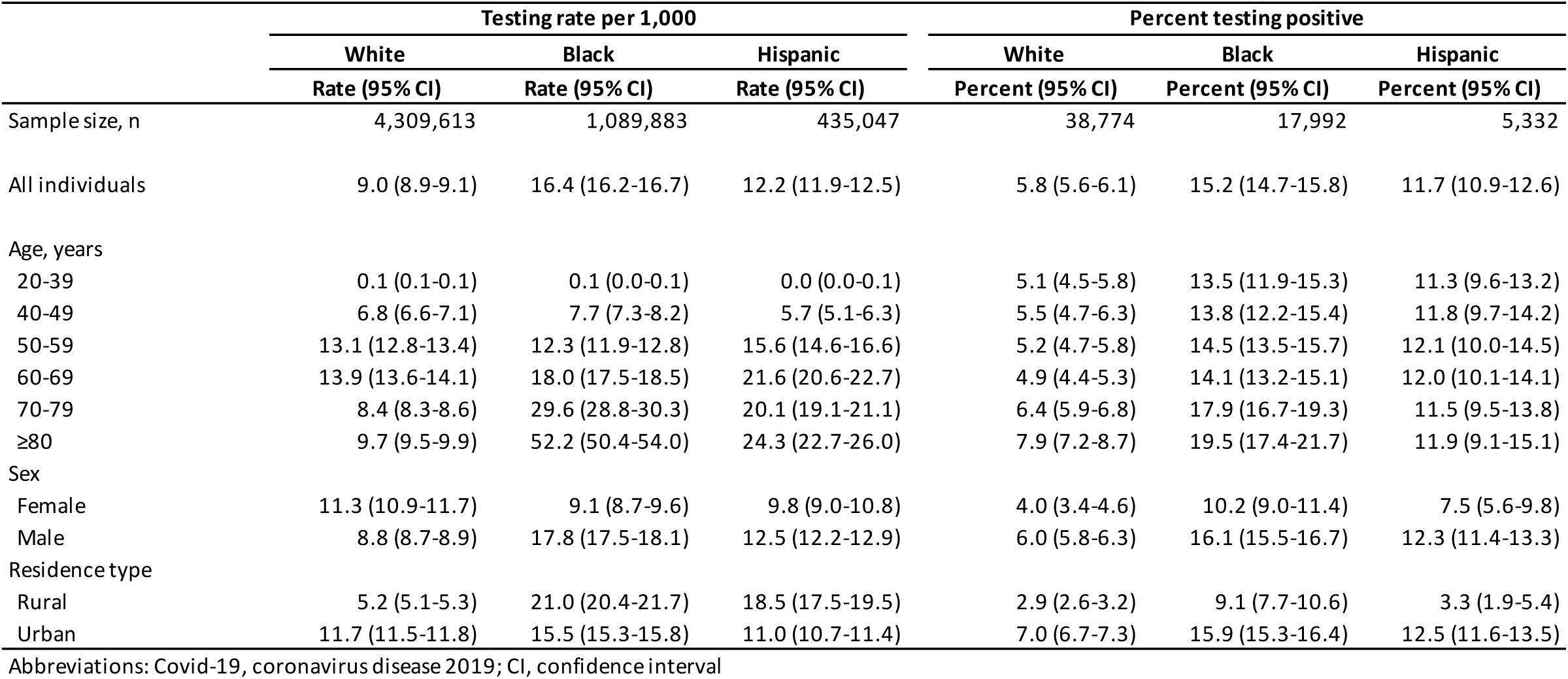
Covid-19 testing by race/ethnicity among all individuals in care as of May 4, 2020

Among individuals tested for Covid-19, the proportion with a positive test varied by race/ethnicity (**Table 2**); 5.8% (95% CI 5.6-6.1%) of white, 15.2% (95% CI 14.7-15.8%) of black, and 11.7% (95% CI 10.9-12.6%) of Hispanic individuals tested positive for Covid-19. For all race/ethnicity groups, positive results were highest at older ages, among men, and those residing in urban settings, apart from Hispanic individuals whereby positive results were fairly similar across all ages.

### Multivariable Models of Testing Positive

In age-adjusted analyses (**Table 3**), black race (OR 3.13, 95% CI 2.94-3.33), Hispanic ethnicity (2.25, 95% CI 2.04-2.49), male sex (OR 1.49, 95% CI 1.34-1.65), and urban residence (OR 3.06, 95% CI 2.77-3.38) were associated with increased likelihood of testing positive for Covid-19. These associations remained, although slightly attenuated, in fully adjusted models and models additionally conditioned on site of care. In models accounting for a wide range of demographic and clinical risk factors and conditioned on site of care, black (OR 1.96, 95% CI 1.81-2.12) and Hispanic (OR 1.73, 95% CI 1.53-1.96) individuals remained strongly associated with increased likelihood of testing positive for Covid-19 (**Figure 2a**). Independent of race/ethnicity and all other adjusting factors, men were more likely to test positive than women (OR 1.74, 95% CI 1.55-1.96) and those living in urban settings were more likely than to test positive than those living in rural settings (OR 1.38, 95% CI 1.23-1.55).

**Figure 2.**
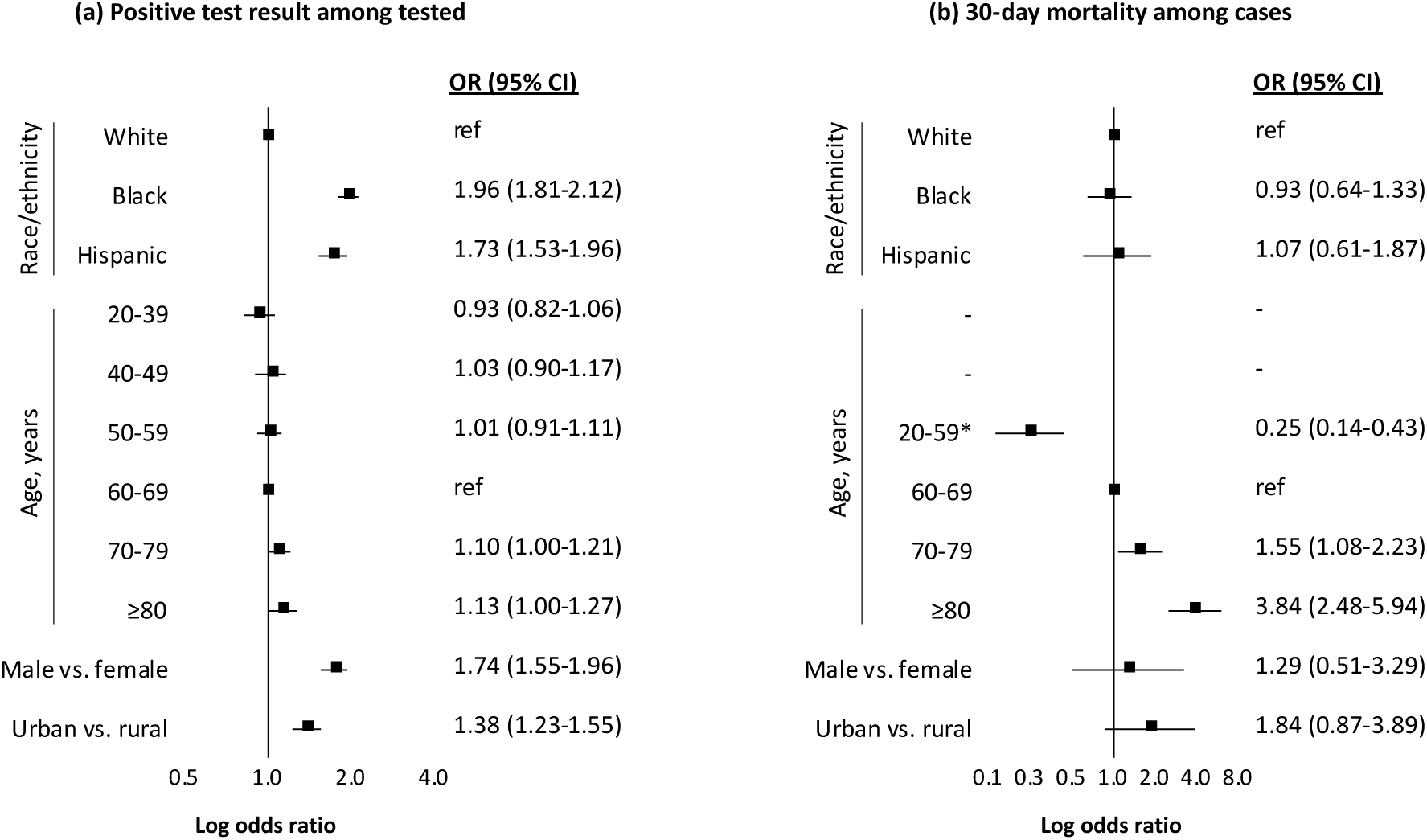
Adjusted associations with testing positive for Covid-19 and subsequent 30-day mortality as of May 4, 2020 Abbreviations: OR, odds ratio; CI, confidence interval. Notes: Both models conditioned on site of care and adjusted for baseline comorbidity (asthma, cancer, chronic kidney disease, chronic obstructive pulmonary disease, diabetes mellitus, hypertension, liver disease, vascular disease), substance use (alcohol consumption, alcohol use disorder, smoking status), and medication history (angiotensin converting enzyme inhibitor, angiotensin II receptor blocker). *Low number of mortality events in age groups 20-39 (0 events) and 40-49 (3 events) thus grouped with 50-59 (23 deaths).

**Table 3.** Associations with testing positive and subsequent 30-day mortality, February 8 to May 4, 2020

|  | Positive test result among tested |  |  | 30-day mortality among cases* |  |  |
| --- | --- | --- | --- | --- | --- | --- |
|  | Age-adjusted OR (95% CI) | Multivariable OR<sup>a</sup> (95% CI) | Conditional OR<sup>b</sup> (95% CI) | Age-adjusted OR (95% CI) | Multivariable OR<sup>a</sup> (95% CI) | Conditional OR<sup>b</sup> (95% CI) |
| Number complete cases | 62,098 | 57,071 | 52,906 | 2,420 | 2,420 | 2,391 |
| Number of events | 5,630 | 5,145 | 5,086 | 284 | 284 | 280 |
| Race/ethnicity |  |  |  |  |  |  |
| White | ref | ref | ref | ref | ref | ref |
| Black | 3.13 (2.94-3.33) | 2.83 (2.65-3.03) | 1.96 (1.81-2.12) | 1.10 (0.83-1.46) | 0.93 (0.68-1.25) | 0.93 (0.64-1.33) |
| Hispanic | 2.25 (2.04-2.49) | 1.80 (1.63-1.99) | 1.73 (1.53-1.96) | 1.14 (0.71-1.85) | 1.05 (0.64-1.72) | 1.07 (0.61-1.87) |
| Age, years |  |  |  |  |  |  |
| 20-39 | 0.90 (0.81-1.00) | 0.94 (0.83-1.06) | 0.93 (0.82-1.06) |  |  |  |
| 40-49 | 1.01 (0.90-1.12) | 0.96 (0.86-1.08) | 1.03 (0.90-1.17) |  |  |  |
| 50-59 | 1.07 (0.98-1.17) | 0.99 (0.90-1.09) | 1.01 (0.91-1.11) | 0.19 (0.11-0.31) | 0.27 (0.16-0.46) | 0.25 (0.14-0.43) |
| 60-69 | ref | ref | ref | ref | ref | ref |
| 70-79 | 1.04 (0.96-1.13) | 1.16 (1.07-1.27) | 1.10 (1.00-1.21) | 2.01 (1.45-2.80) | 1.80 (1.28-2.54) | 1.55 (1.08-2.23) |
| ≥80 | 1.20 (1.09-1.33) | 1.25 (1.12-1.40) | 1.13 (1.00-1.27) | 5.52 (3.79-8.02) | 4.62 (3.07-6.94) | 3.84 (2.48-5.94) |
| Sex, male vs. female | 1.49 (1.34-1.65) | 1.87 (1.68-2.08) | 1.74 (1.55-1.96) | 1.34 (0.56-3.20) | 1.21 (0.50-2.94) | 1.29 (0.51-3.29) |
| Residence type, urban vs. rural | 3.06 (2.77-3.38) | 2.35 (2.13-2.61) | 1.38 (1.23-1.55) | 2.01 (1.10-3.65) | 1.90 (1.03-3.53) | 1.84 (0.87-3.89) |

Abbreviations: Covid-19, coronavirus disease 2019; OR, odds ratio; CI, confidence interval
\*Models of 30-day mortality limited to cases testing positive for Covid-19 on or before April 3, 2020 to allow 30 days of follow-up
<sup>a</sup>Additionally adjusted for baseline comorbidity (asthma, cancer, chronic kidney disease, chronic obstructive pulmonary disease, diabetes mellitus, hypertension, liver disease, vascular
<sup>b</sup>Modeling stations with five or more Covid-19 cases and conditioning on VA site of care
<sup>c</sup>Low number of mortality events in age groups 20-39 (0 events) and 40-49 (3 events) thus grouped with 50-59 (23 deaths)

### Multivariable Models of 30-day Mortality

There were 2,420 individuals who tested positive for Covid-19 on or before April 3, 2020, of whom 284 died within 30 days. Most deaths (n=228, 80.3%) occurred in hospital. In age-adjusted, multivariable, and conditional analyses (**Table 3**), black race (OR 0.93, 95% CI 0.64-1.33) and Hispanic ethnicity (OR 1.07, 95% CI 0.61-1.87) were not associated with 30-day mortality (**Figure 2b**). Urban residence was associated with 30-day mortality in an age-adjusted model (OR 2.01, 95% CI 1.10-3.65), and this association remained elevated but lost significance once accounting for other demographic, clinical, and behavioral factors (OR 1.84, 95% CI 0.87-3.89). There was no association between sex and 30-day mortality in any model, but confidence intervals were wide likely due to the low number of women.

### Sensitivity analyses

Associations with testing positive for Covid-19 were broadly similar over time, but there were some important distinctions. While the association between Hispanic ethnicity and testing positive did not change over time, the association with black race attenuated in the most recent 30 days (OR 2.59, 95% CI 2.29-2.94 between February 8 to April 3, 2020; OR 1.64, 95% CI 1.48-1.82 between April 4 to May 4, 2020, **Supplementary Table S2**). Results from a model of 30-day mortality restricted to inpatient deaths did not alter conclusions from primary analyses (OR 1.19, 95% CI 0.89-1.59 for black race; OR 0.96, 95% CI 0.61-1.52 for Hispanic ethnicity; data not otherwise shown).

## Discussion

This study is the largest in the world to examine racial and ethnic disparities in testing and subsequent Covid-19 mortality representing approximately 6 million individuals receiving care in the US. We found that black and Hispanic individuals were more likely to be tested and test positive for Covid-19 than white individuals, even after accounting for underlying health conditions, other demographics, and geographic location. While individuals from minority backgrounds appeared to experience excess burden of Covid-19, among those infected, there was no observed difference in 30-day mortality by race/ethnicity group.

### Key strengths and limitations

This study was the first to elucidate racial disparities in testing patterns of Covid-19 independent of underlying health status and other key factors. Key strengths of this study were that it was based on well annotated national electronic health record data from a team with decades of experience using VA data, enabling a rapid and reliable analysis of Covid-19 by race and ethnicity. This analysis utilized patients’ records from an entire healthcare system, which made it less prone to collider bias (i.e., non-random selection of individuals into a study) than other Covid-19 studies limited to individuals testing positive and admitted to hospital.^17^ Unlike other national healthcare systems, linkage to Covid-19 testing data or outcomes was not required as the integrated nature of VA healthcare provided at over 1200 sites nationally allows all information to be stored in its national Corporate Data Warehouse. We used validated algorithms to accurately extract information on and adjust models for a wide range of clinical, behavioral, and geographic factors with very little missingness in the data. The scale of VA data also allowed us to assess the impact of Covid-19 across separate race/ethnicity groups, which would have masked important differences between black and Hispanic individuals. We continue to monitor numbers for individuals of other minority backgrounds and plan to follow-up these analyses when there are sufficient numbers for analysis.

While this analysis adds information, its limitations must be kept in mind. First, this study was conducted on Veterans currently receiving care in the VA who are older and have a higher prevalence of chronic health conditions and risk behaviors than the general US population.^18-20^ However, prior research has established that after adjusting for age, sex, race/ethnicity, region, and residence type, all of which were included in this study, there is no difference between total disease burden between Veterans and non-Veterans.^20^ Therefore, associations reported in this study are likely generalizable to the wider US population. Second, while individuals in VA care represent a diversity of backgrounds, women represented a small proportion of individuals in the sample. Thus, we were not powered to assess interactions between sex and race/ethnicity. Third, beyond adjusting for rural/urban and site of testing, we were not able to explore likely causes of the pronounced differential burden of Covid-19 among minority individuals. More detailed information on nursing home residence, socioeconomic status (e.g., type of employment, income, number of individuals in household) were unavailable or not consistently recorded in VA data as in most other electronic health record data sources. Fourth, as is true outside the VA, a small proportion of individuals have been tested (~1%) and rates of testing vary widely by site and within important subgroups. However, while initial testing was limited, by mid-April the VA began testing all individuals admitted to hospital and before any inpatient or outpatient procedures are done for clearance even in those not suspected to have Covid-19. We performed sensitivity analyses to see if associations with testing positive changed over time and found conclusions were broadly similar but with some important distinctions. However, our models for testing positive should not be interpreted as a proxy of likelihood of infection since those with mild symptoms were unlikely to have received testing, particularly in the early stages of the outbreak.

### Findings in context

Our findings of racial and ethnic disparities in Covid-19 provide important distinctions from previous reports in the US and other countries with ethnically diverse populations. To our knowledge, the largest national study of race/ethnicity and Covid-19 in the US to date included 580 individuals across 14 states, showing that black individuals represented 33% of Covid-19 hospitalizations despite only making up 18% of the total population studied.^7^ In the UK, which was the first country with an ethnically diverse population to experience a Covid-19 outbreak,^21^ a study of 17 million individuals showed that those from minority backgrounds had a substantially higher risk of in-hospital death from Covid-19, which was not fully explained by underlying health conditions or deprivation.^22^ While our study also found racial and ethnic disparities that were not fully explained by health status, we found that these disparities occurred primarily at stages prior to hospitalization (i.e., testing positive for Covid-19). We found no evidence of racial or ethnic disparities in 30-day mortality once models were restricted to those who tested positive for Covid-19.

We demonstrated that black and Hispanic individuals were more likely to test positive than their white counterparts even after accounting for underlying health conditions, other demographics, residence type, and site of care. Our finding may still be an underestimate of the US population risk as health disparities in VA tend to be smaller than in the private sector.^23^ Based on experience with 1918 Spanish Flu and the 2009 H1N1 epidemic, public health experts have warned that racial and ethnic minority populations may be at higher risk during infectious disease outbreaks due to underlying health conditions, lower access to care, and socioeconomic conditions.^24 25^ Prior reports have highlighted that members of racial and ethnic minorities are more likely to live in densely populated areas or multi-generational households, and minority groups are overrepresented in jails, prisons, and detention centers, all of which lead to reduced capacity to implement physical distancing.^26-30^ Similarly, black and Hispanic workers are more likely than their white counterparts to be workers in essential industries who continue to work outside the home despite outbreaks in their communities making them more prone to infection.^30-32^

Testing rates for Covid-19 in the VA were higher among black and Hispanic individuals compared to white individuals. Local reporting from metropolitan areas with large minority populations, including New York City^33^ and Chicago,^34^ have highlighted the disproportionate impact of Covid-19 in minority communities. We showed that VA facilities in these cities and others around the country that conducted the highest number of Covid-19 tests also had the highest proportion of black individuals in care. There was a striking difference in the rate of Covid-19 testing by age across race/ethnicity groups, whereby higher rates of testing were observed for middle-aged, white individuals while testing rates steadily increased with age and was highest among those aged 80 or more years among black and Hispanic individuals. White individuals living in urban residences were more likely to be tested than those living in rural residences while the opposite was true among black and Hispanic individuals. Among all individuals tested, those living in urban residences were more likely to test positive for Covid-19 than those living in rural residences. Finally, women were more likely to be tested for Covid-19 than men, but men were twice as likely to test positive. This association strengthened after adjustment and in conditional analyses but should be considered preliminary given limited numbers of women in this analysis.

### Policy implications

We have been able to confirm several local and national reports that black and Hispanic individuals in the US are impacted by Covid-19 differentially to their white counterparts. While other studies have focused on hospitalized individuals, this study is the first and largest to show that, at least through May 4, 2020, black and Hispanic patients were more likely to be tested and to test positive. We found no difference in 30-day mortality among Covid-19 cases. These findings highlight the urgent need for improved strategies to contain and prevent further outbreaks in the US, particularly within black and Hispanic communities.

### Future research

We appeal to other researchers investigating racial and ethnic disparities to perform analyses on the entire population at-risk for Covid-19, where data are available, and compare findings associated at each stage in the clinical course of Covid-19, from testing to outcomes. In this paper, we focused only on 30-day mortality among Covid-19 cases. We plan to explore other outcomes, including hospitalization, intensive care, and intubation, in subsequent analyses to examine whether racial and ethnic disparities exist in the clinical course of Covid-19 after testing positive and before death. Among other factors, future research should consider the role of employment type, nursing home residence, and incarceration. Other racial and ethnic minorities in the US deserve attention and while we did not have enough power to include other groups in this analysis, we will continue to monitor these numbers for future research.

## Conclusion

While minority individuals with Covid-19 did not appear to have worse outcomes, black race and Hispanic ethnicity were strongly associated with testing positive for Covid-19 suggesting a substantial excess burden of Covid-19 in US minority populations.

## Data Availability

The analytic data sets used are not permitted to leave the VA firewall. This limitation is consistent with the authors' past work and with other studies based on VA data.

## Acknowledgements

The views and opinions expressed in this manuscript are those of the authors and do not necessarily represent those of the Department of Veterans Affairs or the United States Government. The authors wish to recognize Dr. Kendall Bryant as the NIAAA Scientific Collaborator.

## Contributorship

CR and AJ conceived and led the project overall and are guarantors. Contributions are as follows: Data curation CR, FK, JT, LP, JK, MS, RH, MH; Formal analysis CR; Funding acquisition AJ; Methodology CR, JT, and AJ led with support from LP, RH, VL, KA, KC, TT, MF; Project administration CR and AJ; Visualization CR, JT, LP, CJ; Writing (original draft) CR and AJ; All authors reviewed, edited, and approved the final manuscript.

## Appendix

**Table S1.** International Classification of Diseases, Tenth Revision, Clinical Modification (ICD-10-CM) Diagnosis Codes

| ICD-10-CM codes |  |
| --- | --- |
| **Comorbid conditions** |  |
| **Asthma** | J45.X |
| **Cancer** |  |
| Cancer | C00.X-C43.X, C45.X-C76.X, C80.X-C96.X, C7A.X |
| Metastatic cancer | C77.X-C79.X |
| **Chronic obstructive pulmonary disease** | J41.X, J42.X, J43.X, J44.X |
| **Chronic kidney disease** | I12.0X, I13.1X, N03.2X-N03.7X, N18.X, N19.X, N05.2X-N05.7X, N25.0X, Z49.0X - Z49.2X, Z94.0X, Z99.2X |
| **Diabetes mellitus** | E08.X, E10.X, E11.X, E13.X |
| **Hypertension** | I10.X-I13.X, I15.X, I16.X |
| **Liver disease** |  |
| Hepatitis B virus | B16.X, B18.0X, B18.1X, B19.1X, Z22.51 |
| Hepatitis C virus | B17.10, B17.11, B18.2, B19.20, B19.21, Z22.52 |
| Hepatic decompensation | I85.01, K65.2, K70.31, K72.1X, K72.9X, K76.7, R18.8 |
| Other mild liver disease | B18.8X, B18.9X, K70.0X-K70.2X, K70.30, K70.9X, K71.3X-K71.5X, K71.7X, K73.X, K74.X, K76.0X, K76.2X-K76.4X, K76.8X, K76.9X, Z94.4 |
| Other severe liver disease | K76.6, I85.00, I85.9X, I86.4, I98.2X, K70.4X, K71.1X, K76.5X |
| **Vascular disease** |  |
| Acute myocardial infarction | I21.X (not including I21.AX), I22.X |
| Cardiomyopathy | I42.X, I43.X |
| Coronary heart disease | I20.X, I24.X, I25.10, I25.110, I25.2, I25.3, I25.41, I25.42, I25.5, I25.700, I25.710, I25.720, I25.730, I25.750, I25.760, I25.790, I25.8X, I25.9 |
| Heart failure | I09.9, I11.0, I25.5, I13.0, I13.2, I50.X, P29.0 |
| Cerebrovascular accident | I60.X-I69.X, G45.X, G46.X, H34.0 |
| Peripheral vascular disease | I70.X, I71.X, I73.1-I73.9, I77.1, I79.0, I79.2, K55.1X, K55.8X, K55.9X, Z95.8X, Z95.9 |
| **Substance use** |  |
| **Alcohol use disorder** | F10.1X, F10.2X |

**Table S2.** Associations with testing positive for Covid-19 over time

|  | Positive test result among tested |  |
| --- | --- | --- |
|  | February 8 to April 3 | April 4 to May 4 |
| Number complete cases | 14,813 | 38,093 |
| Number of events | 2,391 | 2,695 |
| Race/ethnicity |  |  |
| White | ref | ref |
| Black | 2.59 (2.29-2.94) | 1.64 (1.48-1.82) |
| Hispanic | 1.77 (1.46-2.14) | 1.71 (1.45-2.00) |
| Age, years |  |  |
| 20-39 | 0.80 (0.65-0.97) | 0.94 (0.79-1.12) |
| 40-49 | 0.79 (0.65-0.96) | 1.10 (0.92-1.31) |
| 50-59 | 0.87 (0.74-1.02) | 1.10 (0.96-1.25) |
| 60-69 | ref | ref |
| 70-79 | 1.10 (0.95-1.29) | 1.11 (0.98-1.26) |
| ≥80 | 1.02 (0.82-1.26) | 1.26 (1.08-1.47) |
| Sex, male vs. female | 2.31 (1.92-2.78) | 1.42 (1.22-1.66) |
| Residence type, urban vs. rural | 1.26 (1.05-1.52) | 1.41 (1.22-1.64) |

Abbreviations: Covid-19, coronavirus disease 2019; OR, odds ratio; CI, confidence interval
Notes: OR (95% CI) from conditional logistic regression modeling stations with five or more Covid-19 cases and conditioning on VA site of care. Additionally adjusted for baseline comorbidity (asthma, cancer, chronic kidney disease, chronic obstructive pulmonary disease, diabetes mellitus, hypertension, liver disease, vascular disease), substance use (alcohol consumption, alcohol use disorder, smoking status), and medication history (angiotensin converting enzyme inhibitor, angiotensin II receptor blocker)
<sup>a</sup>Low number of mortality events in age groups 20-39 (0 events) and 40-49 (3 events) thus grouped with 50-59 (23 deaths)

